# Development and Validation of a Two-Stage NLP-LLM System for Automated Extraction of Deprescribing Recommendations from Discharge Summaries

**DOI:** 10.64898/2026.04.29.26352010

**Authors:** Kenji Fujita, Marion Matheson, Bhavya Valecha, Sarah N Hilmer

## Abstract

**Introduction:** Polypharmacy in older adults is associated with increased risks of adverse drug events and functional decline. Discharge summaries often contain deprescribing recommendations, but these are frequently overlooked due to documentation complexity.

**Objective:** To develop and validate a two-stage hybrid system combining rule-based natural language processing (NLP) and large language model (LLM) for automated extraction of deprescribing recommendations from discharge summaries.

**Methods:** This retrospective cohort study included 850 discharge summaries from patients aged ≥65 years with hospitalisation ≥48 hours across six public hospitals in New South Wales, Australia. Model 1 (rule-based NLP) extracted discharge medications and candidate sentences containing pre-defined deprescribing keywords. Model 2 (open-source LLM) classified candidate sentences into five categories. Data were split into training (80%) and test (20%) sets. Gold standard classifications were established by independent reviews, followed by adjudication of discrepancies.

**Results:** Model 1 extracted 9,631 discharge medications (median 11 per patient) and 1,061 candidate sentences from 850 patients (median age 82.8 years). Model 2 achieved an F1 score of 0.91 and accuracy of 0.90 on the test set. Inter-rater reliability showed substantial agreement (Cohen’s kappa = 0.70). The most frequently identified medications recommended for deprescribing were antibiotics and opioids. The most common misclassification was incorrectly identifying actions completed during hospitalisation as post-discharge recommendations. The combined processing time averaged 12.6 seconds per discharge summary.

**Conclusions:** A two-stage hybrid approach combining rule-based NLP and open-source LLM can accurately extract deprescribing recommendations from discharge summaries, enabling cost-efficient, privacy-compliant local deployment.

**Key Points:**

- A two-stage system combining rule-based NLP and open-source LLM extracted and classified deprescribing recommendations from 850 discharge summaries, achieving an F1 score of 0.91 and accuracy of 0.90.
- The use of an open-source LLM (Llama 3.3) enables cost-efficient, privacy-compliant local deployment in healthcare institutions.
- Antibiotics and opioids were the most frequently identified medications recommended for deprescribing in discharge summaries.

## 1 Introduction

Polypharmacy, commonly defined as the concurrent use of five or more medications, is associated with increased risks of adverse drug events, medication non-adherence, and functional decline.[1, 2] Deprescribing, which aims to reduce polypharmacy, is a process of withdrawing an inappropriate medication (one in which the risks outweigh the benefits in the individual, which include high risk and unnecessary medications), with the goal of improving patient outcomes.[3] Internationally, approximately 84% of older patients would be willing to stop one or more of their medications if recommended by the prescriber.[4] Therefore, deprescribing supported by a health care professional is important in older adults.[3]

The hospital discharge transition represents a critical period for deprescribing, as medication regimens frequently change and errors occur after discharge.[5, 6] While discharge summaries often contain deprescribing recommendations,[7] their length and complexity may limit effective communication to primary care providers.[8, 9]

Understanding what deprescribing recommendations are documented in discharge summaries is important for quality improvement and research. Traditional manual review for extracting such recommendations is labour-intensive, time-consuming, and susceptible to human error. Recent advances in artificial intelligence, particularly natural language processing (NLP) and large language models (LLMs), offer promising solutions to automate this process.[10, 11] A hybrid approach combining rule-based NLP for efficient keyword identification with LLMs for contextual understanding has shown potential for clinical information extraction.[12] Recent studies have demonstrated that prompt engineering strategies, including few-shot learning, can enhance LLM performance for clinical named entity recognition and related information extraction tasks.[13]

While several studies have explored automated identification of potentially inappropriate medications using established criteria such as STOPP (Screening Tool of Older Persons’ Prescriptions) criteria, research focusing on extraction of actual deprescribing recommendations documented by hospital clinicians remains limited.[14, 15] Therefore, this study aimed to develop and validate a two-stage hybrid system combining rule-based NLP (Model 1) for identifying candidate sentences and extracting medication lists, with an open-source LLM to classify actual deprescribing recommendations.

## 2 Methods

### 2.1 Study design and data source

This retrospective cohort study used discharge summaries from six public hospitals in New South Wales, Australia. All hospitals used the same electronic medical records platform (Cerner EMR, Oracle Health). Patients aged ≥65 years with hospitalisation ≥48 hours discharged between January 4 and March 28, 2022 were included (Figure 1). Exclusion criteria were in-hospital death and absence of discharge medication documentation. A total of 900 discharge summaries were randomly sampled. After excluding 50 discharge summaries (35 deaths, 15 without medications), 850 summaries were analysed. Ethics approval was granted by the Northern Sydney Local Health District Human Research Ethics Committee (2021/ETH11776).

**Figure 1.**
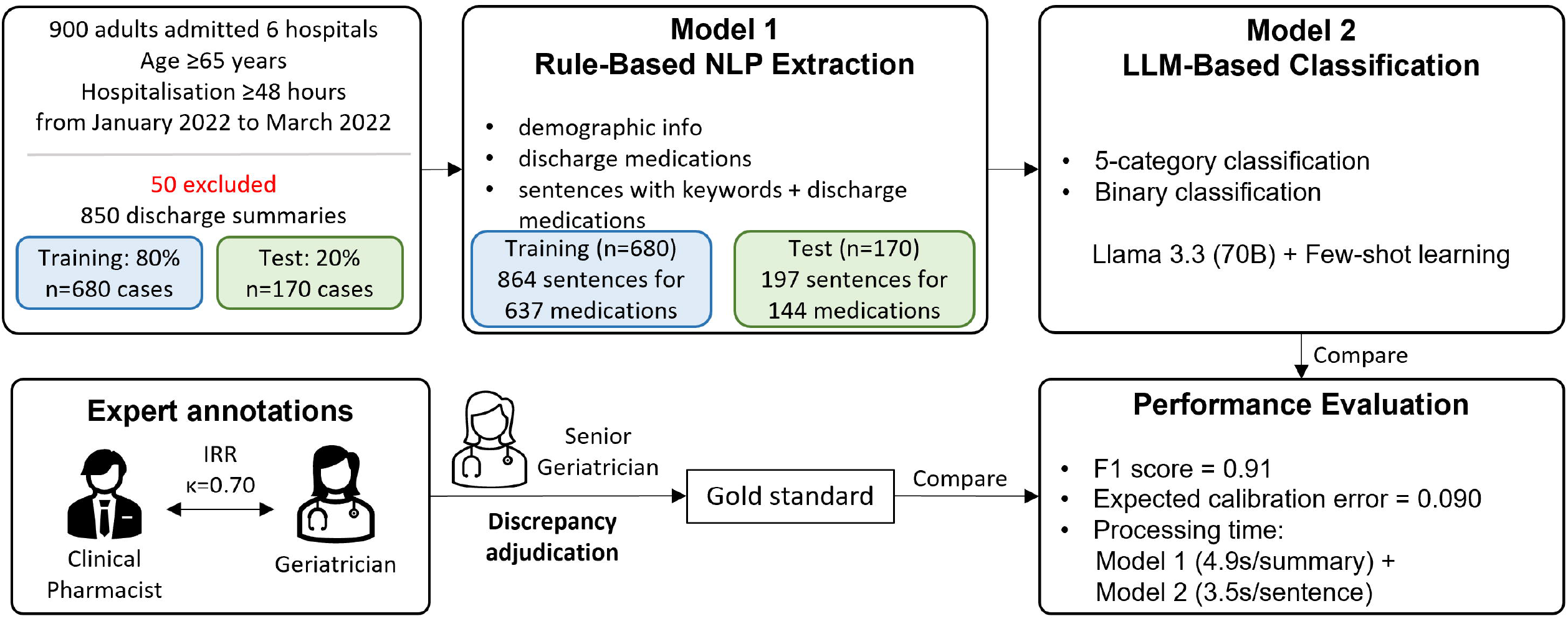
Overview of the two-stage NLP-LLM system development and validation

### 2.2 Development of a two-stage extraction and classification system

Preliminary examination revealed that while patient demographics, admission information, and discharge medications were consistently documented in structured formats across all hospitals, deprescribing recommendations varied considerably in location and format.[16, 17] This variability necessitated a two-stage approach for extraction: Model 1 (rule-based approach) extracted discharge medication lists and candidate sentences for deprescribing recommendations, and Model 2 (LLM-based approach) classified their content (Figure 1).

#### 2.2.1 Model 1: Rule-Based Medication Extraction

Model 1 was designed to extract patient attributes, discharge medication lists, and candidate sentences likely to contain deprescribing recommendations from discharge summaries. Discharge medications were extracted by pattern matching from the “Medication being taken on discharge” section, where medications were listed using active ingredient names (and occasionally brand names).[18] However, when documenting deprescribing recommendations, clinicians exhibited considerable variability in how they referred to medications. Therefore, to extract medications as comprehensively as possible, the pattern matching was expanded to multiple medication classification systems. The strategy included active ingredient names and brand names listed in MIMS Australia,[18] drug classes from the WHO Anatomical Therapeutic Chemical (ATC) Classification System (e.g., opioids, antipsychotics),[19] and Therapeutic categories from the Therapeutic Category of Drugs in Japan (e.g., bronchodilator, analgesic).[20] When other abbreviations (e.g., “abx” for antibiotics, “pred” for prednisolone, “vit D” for colecalciferol) not contained in these systems appeared in discharge summaries, those terms were added to the pattern-matching list.

To correct spelling errors of medication names, a similarity assessment was implemented based on the Levenshtein distance and a derived similarity score. The Levenshtein distance measures the minimum number of single-character edits (insertions, deletions, or substitutions) required to transform one string into another. The similarity score was calculated as follows: Similarity score = (1 − (Levenshtein distance / max(len(string1), len(string2)))) × 100.[21] Optimal thresholds for Levenshtein distance and similarity score were determined through iterative evaluation on the training dataset. This algorithm enabled matching of morphological variations between medication classification systems and discharge summary text.

From the entire discharge summary, the model extracted sentences containing pre-defined keywords suggesting deprescribing. This keyword list was created through an iterative evaluation process using the training data. The initial list included explicit deprescribing terms such as stop, cease, down-titrate, reduce, wean, taper, and discontinue, but was expanded through evaluation of discharge summaries. After extracting sentences with keywords, the model evaluated whether each sentence contained the discharge medication names, and sentences with both keywords and medication names were identified as candidate sentences that contains deprescribing recommendations for subsequent steps. Model 1 generated a data sheet containing patient demographic, basic admission information, discharge medication list, and deprescribing candidate sentences with associated medication names.

#### 2.2.2 Model 2: LLM-Based Classification

Model 2 classified candidate sentences into five categories: 1) no deprescribing recommendation, 2) dose adjustment with unclear direction (unclear if up or down titration), 3) dose reduction without aim to cease, 4) dose reduction with aim to cease, and 5) cessation. To maximise classification accuracy by the LLM, a few-shot learning strategy was implemented (Supplementary File 1). To ensure reproducibility, the temperature parameter was set to 0. Llama 3.3 (70 billion parameters) was used in a computing environment consisting of Windows 11, Intel Core i9-12900 CPU, NVIDIA RTX 6000 Ada Generation GPU, 128GB RAM, and Python version 3.10.11.

### 2.3 Model development and validation

The 850 discharge summaries were randomly divided into 80% (680 cases) training data and 20% (170 cases) test data. Using the training data, iterative optimisation processes were implemented including selection and refinement of pre-defined keywords for deprescribing recommendations, identification of the abbreviation/alternative terminology for medications, threshold setting for typographical error correction (i.e., the Levenshtein distance and similarity score) and prompt engineering. This process continued until the model achieved an F1 score >0.9 on the training data. The finalised model was then evaluated on the remaining test data.

### 2.4 Data analysis

The primary outcome was the binary classification performance of Model 2 in identifying deprescribing recommendations (present vs. absent) on the test set, evaluated using sensitivity, specificity, positive predictive value (PPV), negative predictive value (NPV), F1 score, and accuracy. Classification performance was also evaluated separately for each of the six hospitals. When multiple candidate sentences mentioned the same medication in a discharge summary, the final classification was determined using a hierarchical priority rule based on the strength of deprescribing recommendation from Category 1: cessation (highest priority) >> Category 5: no deprescribing recommendation (lowest priority). As a sensitivity analysis, performance was also evaluated at the sentence level, where each candidate sentence was independently assessed.

To establish the gold standard for model evaluation, a clinical pharmacist and geriatric trainee independently classified each candidate sentence extracted by Model 1 into the five predefined categories. Inter-rater reliability was assessed using percentage agreement for each of the five categories and Cohen’s kappa coefficient for binary classification. Discrepancies were adjudicated by a senior geriatrician and clinical pharmacologist to determine the final classification used as the gold standard.

For the primary analysis, the five-category classification was converted to binary classification. Categories 3 (dose reduction without aim to cease), 4 (dose reduction with aim to cease), and 5 (cessation) were defined as “deprescribing recommendation present” (coded as 1), while Categories 1 (no deprescribing recommendation) and 2 (dose adjustment with unclear direction) were defined as “deprescribing recommendation absent” (coded as 0). While acknowledging conflict in the literature as to whether dose reduction without the aim of cessation is included in the definition of deprescribing,[22] this binary classification was used for both inter-rater reliability assessment as well as primary outcome.

To evaluate prediction reliability, a consistency-based confidence assessment was implemented. Each candidate sentence was classified five times with a higher temperature setting (T=0.8) to introduce response variability. The final classification was determined through confidence-weighted majority voting, where each prediction was weighted by its self-reported confidence score (0-100). Agreement scores and Expected Calibration Error (ECE) were calculated to assess prediction consistency and confidence calibration.[23]

The following additional analyses were performed: 1) error pattern analysis to identify causes of false positives and false negatives, 2) frequency analysis of predefined keywords used in deprescribing recommendations, 3) identification of the most frequently recommended medications for deprescribing, 4) frequency of spelling errors detected in medication names, and 5) processing time measurement for both models. All medications were classified using ATC codes. All analyses were performed using Python 3.10.11.

## 3 Results

### 3.1 Model 1: Rule-based extraction

Model 1 extracted patient demographic and admission information from 850 discharge summaries across six hospitals (Table 1).

**Table 1.** Patient demographic and clinical characteristics across six hospitals.

|  | Hospital A<br>(n=164) | Hospital B<br>(n=163) | Hospital C<br>(n=159) | Hospital D<br>(n=167) | Hospital E<br>(n=27) | Hospital F<br>(n=170) |
| --- | --- | --- | --- | --- | --- | --- |
| Age, mean (SD), years | 81.6 (8.5) | 81.3 (8.1) | 83.5 (9.3) | 83.2 (8.3) | 82.6 (8.9) | 81.7 (8.3) |
| Female, % | 56.7% | 50.9% | 47.8% | 56.9% | 48.1% | 61.8% |
| Length of stay, median [IQR], days | 6.9 [4.2-11.4] | 6.6 [3.9-14.5] | 6.1 [4.0-10.6] | 6.9 [4.1-12.7] | 24.9 [13.3-52.9] | 8.5 [4.8-27.9] |
| Charlson Comorbidity Index, median [IQR] | 1.0 [0.0-2.0] | 1.0 [0.0-2.0] | 1.0 [0.0-2.0] | 1.0 [0.0-2.0] | 0.0 [0.0-1.5] | 1.0 [0.0-2.0] |
| Interpreter required, % | 1.8% | 9.2% | 6.9% | 15.6% | 7.4% | 0.6% |
| Born in Australia, % | 79.9% | 55.2% | 52.2% | 48.5% | 66.7% | 76.5% |
| Number of medications, median [IQR] | 11.0 [7.0-14.0] | 11.0 [7.0-15.0] | 10.0 [7.0-14.0] | 10.0 [7.0-15.0] | 12.0 [9.0-16.0] | 12.0 [8.0-16.0] |
| Five most frequent medical services | Geriatrics:<br>82 (50.0%)<br>Orthopaedic<br>Surgery:<br>16 (9.8%)<br>Neurology:<br>12 (7.3%)<br>General Surgery:<br>10 (6.1%)<br>Respiratory<br>Medicine:<br>6 (3.7%) | Medicine:<br>62 (38.0%)<br>Respiratory<br>Medicine:<br>25 (15.3%)<br>General Surgery:<br>18 (11.0%)<br>Geriatrics:<br>11 (6.7%)<br>Orthopaedic<br>Surgery:<br>8 (4.9%) | Geriatrics:<br>79 (49.7%)<br>Orthopaedic<br>Surgery:<br>13 (8.2%)<br>Cardiology:<br>9 (5.7%)<br>Neurology:<br>9 (5.7%)<br>Respiratory<br>Medicine:<br>7 (4.4%) | Medicine:<br>113 (67.7%)<br>General Surgery:<br>19 (11.4%)<br>Orthopaedic<br>Surgery:<br>15 (9.0%)<br>Cardiology:<br>14 (8.4%)<br>Rehabilitation:<br>3 (1.8%) | Geriatrics:<br>22 (81.5%)<br>Rehabilitation:<br>5 (18.5%) | Geriatrics:<br>84 (49.4%)<br>Medicine:<br>54 (31.8%)<br>Cardiology:<br>14 (8.2%)<br>General Surgery:<br>8 (4.7%)<br>Neurology:<br>5 (2.9%) |

Abbreviations: SD: standard deviation, IQR: interquartile range

The median age was 82.8 years (IQR: 75.8-88.9), and 54.7% were female. The median length of hospital stay was 7 days (IQR: 4-14). Patients were prescribed a median of 11.0 medications on discharge (IQR: 7.0-15.0). The most common medical services were geriatrics (278 patients, 32.7%), followed by general medicine (235 patients, 27.6%) and general surgery (55 patients, 6.5%). In addition, Model 1 extracted 9,631 discharge medications (7,755 from training set, 1,876 from test set). Through iterative evaluation on the training data, 27 distinct keyword patterns were identified (Supplementary File 1). As a result, Model 1 identified 1,338 sentences containing predefined deprescribing keywords. After medication-sentence linkage, 1,061 candidate sentences were generated (864 from training set, 197 from test set) and forwarded to Model 2. These sentences corresponded to 781 unique discharge medications (637 in training set, 144 in test set). Cohen’s kappa coefficient for inter-rater reliability between the two assessors was 0.704, indicating substantial agreement. Agreement rates across the five categories were: no deprescribing recommendation (71.4%), dose adjustment with unclear direction (93.2%), dose reduction without aim to cease (52.4%), dose reduction with aim to cease (96.5%), and cessation (97.8%).

### 3.2 Model 2: LLM-based classification performance

Model 2 was developed using 637 candidate sentences, corresponding to 637 unique discharge medications after removing within-summary duplicates, from the training set. The classification prompt underwent several iterative refinements, and the finalised version is provided in the Supplementary File 2. For binary classification (deprescribing present vs. absent), the model achieved good performance with an F1 score of 0.91 (Table 2), meeting the predefined performance threshold of an F1 score >0.9.

**Table 2.** Performance of the LLM-based deprescribing classification model in training and test datasets.

| Dataset | TP | TN | FP | FN | Sensitivity | Specificity | PPV | NPV | F1 Score | Accuracy |
| --- | --- | --- | --- | --- | --- | --- | --- | --- | --- | --- |
| Training dataset<br>(n=637 sentences) | 332 | 241 | 55 | 9 | 0.97 | 0.81 | 0.86 | 0.96 | 0.91 | 0.90 |
| Test dataset<br>(n=144 sentences) | 73 | 56 | 11 | 4 | 0.95 | 0.84 | 0.87 | 0.93 | 0.91 | 0.90 |

n=781 candidate deprescribing recommendations for unique medications. Abbreviations: LLM: large language model (Llama 3.3 70B).
Abbreviations: TP: true positive, TN: true negative, FP: false positive, FN: false negative, PPV: positive predictive value, NPV: negative predictive value.

The finalised model was validated on 144 candidate sentences from the test set (170 discharge summaries). For the primary binary classification outcome, the model showed strong performance with an F1 score 0.91 and accuracy 0.90. When the model performance was evaluated separately for each of the six hospitals in the test set, F1 scores varied substantially across hospitals (range: 0.67-0.97, Supplementary File 3).

The confusion matrix for five-category classification at the medication level (n=781 across training and test sets combined) is presented in Table 3, demonstrating varying performance across categories: 1) no deprescribing recommendation 93.0%, 2) dose adjustment with unclear direction 94.5%, 3) dose reduction without aim to cease 30.6%, 4) dose reduction with aim to cease 65.1%, and 5) cessation 88.2%. At the sentence level (n=1,061), classification accuracies ranged from 67.0% (no deprescribing recommendation) to 92.1% (dose adjustment with unclear direction) across the five categories (Supplementary File 4).

**Table 3.** Confusion matrices for five-category deprescribing recommendation classification.

|  |  | Gold Standard |  |  |  |  |
| --- | --- | --- | --- | --- | --- | --- |
|  |  | Cessation | Dose reduction with aim to cease | Dose reduction without aim to cease | Dose adjust (direction unclear) | No deprescribing |
| LLM | Cessation | 247 | 3 | 0 | 0 | 26 |
|  | Dose reduction with aim to cease | 24 | 95 | 5 | 2 | 20 |
|  | Dose reduction without aim to cease | 5 | 11 | 15 | 0 | 18 |
|  | Dose adjust (direction unclear) | 0 | 7 | 1 | 138 | 7 |
|  | No deprescribing | 4 | 1 | 0 | 6 | 146 |

n=781 candidate deprescribing recommendations for unique medications. Abbreviations: LLM: large language model (Llama 3.3 70B). Shaded cells indicate concordance between LLM classification and gold standard.

Consistency analysis demonstrated excellent prediction stability, with 98.5% of cases (769/781) achieving perfect agreement (agreement score = 1.0) across five independent samplings at temperature 0.8. Mean confidence was 94.3% (median: 100.0%). However, calibration analysis revealed moderate miscalibration (ECE = 0.090, Figure 2). The model showed mixed calibration patterns: underconfidence at the 80% confidence level (actual accuracy: 92%) and overconfidence at higher levels (89-100% confidence with 81-91% actual accuracy).

**Figure 2.**
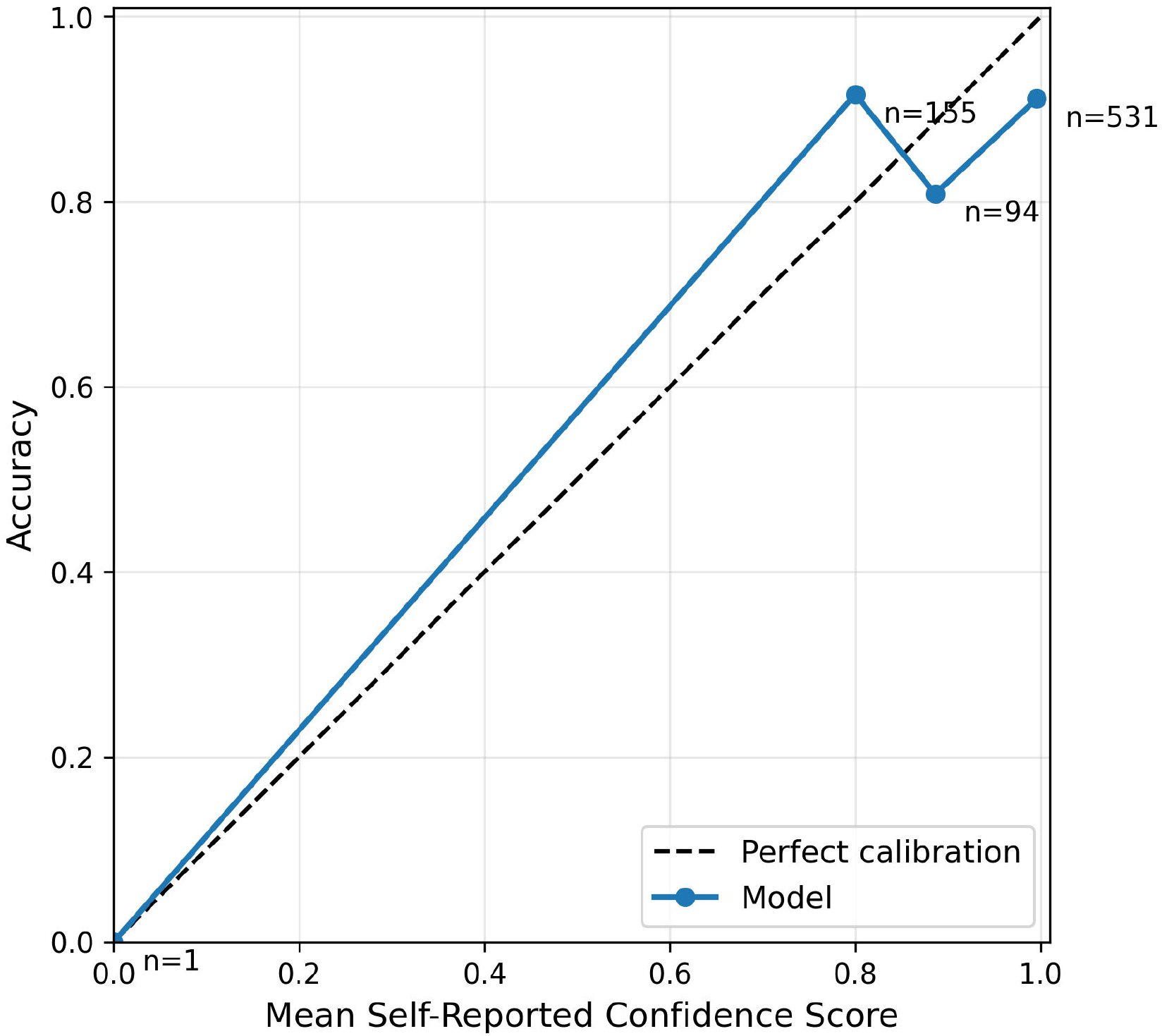
Calibration plot for LLM-based classification. The dashed line represents perfect calibration. Points indicate model performance at each confidence level, with sample sizes shown.

### 3.3 Additional analyses

Across the entire dataset (n=781 medications), 79 misclassifications occurred (10.1% error rate), comprising 63 false positives (8.1%) and 16 false negatives (2.0%, Table 4). Error patterns were consistent between training and test sets. The most common cause of false positive cases was past action misidentified as recommendation (n=23, 36.5%), where actions completed during hospitalisation were misclassified as future recommendations. False negative errors (n=16) resulted entirely from context/meaning misunderstanding, where the model failed to recognise implicit deprescribing recommendations lacking explicit keywords such as “cease”, “stop”, or “wean”.

**Table 4.** Types of misclassification errors.

False Positive Cases (n=63)
| Error Type | Frequency | Definition | Example |
| --- | --- | --- | --- |
| Past action misidentified as recommendation | 23 (36.5%) | Completed actions during hospitalisation were misclassified as future recommendations for the GP. | "Cease telmisartan and reduced dose of sotalol" was misclassified as a deprescribing recommendation for sotalol, but the dose reduction had already been completed during admission. Telmisartan was ceased during the admission. |
| Recommendation for different drug | 17 (27.0%) | A deprescribing recommendation for one medication was incorrectly attributed to a different medication mentioned in the same sentence. | "Symbicort 1 inhalation BD / Prednisolone slow wean every 7 days" contained a weaning recommendation for prednisolone, but was incorrectly attributed to Symbicort (budesonide-formoterol) mentioned in the same sentence. |
| Use of abbreviation or alternative name | 13 (20.6%) | Generic terms, abbreviations, or drug class names in recommendations were not correctly linked to the specific medication being evaluated. | "Please wean opioid analgesia" referred to oxycodone by drug class rather than by name, causing failure to link the recommendation to another analgesic prescribed on discharge. |
| Temporary withholding misidentified as long term cessation | 6 (9.5%) | Instructions to temporarily hold a medication were misclassified as long term deprescribing recommendations. | "Withhold Methotrexate while acutely unwell" indicated temporary cessation during acute illness, not an ongoing, long term deprescribing recommendation. |
| Context or meaning misunderstood | 4 (6.3%) | The model misinterpreted the clinical context, timing, or intent of the recommendation. | "Consider changing pantoprazole to rabeprazole, due to rabeprazole having the least interaction with clopidogrel" was a medication switch recommendation, not deprescribing, but was misclassified as a cessation recommendation for pantoprazole. |

False Negative Cases (n=16)
| Error Type | Frequency | Definition | Example |
| --- | --- | --- | --- |
| Context or meaning misunderstood | 16 (100%) | The model failed to recognise implicit deprescribing recommendations that lacked explicit keywords such as "cease", "stop", or "wean". | "Oral magnesium (Magmin) 1g daily for 5 days, GP to review ongoing need" contained a time-limited prescription with implicit cessation intent, but was classified as no deprescribing recommendation. |

Among all deprescribing recommendations for 418 medications across training and test sets, a total of 454 predefined keywords were used. The most frequently used keyword was “stop” (n=177, 38.8%), followed by “cease” (n=103, 22.6%), and “wean” (n=100, 21.9%), which together accounted for 83.3% of all identified keywords. Supplementary File 5 presents the top 20 medications with deprescribing recommendations. Amoxicillin (including amoxicillin-clavulanate) was the most frequently identified (n=50), with 100% classified as “cessation”. Oxycodone +/-naloxone was the second most frequent (n=42), with the majority (69.0%, 29/42) classified as “dose reduction with aim to cease” rather than immediate cessation (31.0%, 13/42).

The typographical error correction algorithm identified and corrected 9 medication name spelling errors across the dataset: apixiban (2; corrected to *apixaban*), augementin (1; *Augmentin*), cefazolin (1; *cephazolin*), ceflexin (1; *cephalexin*), Cholecalciferol (3; *colecalciferol*), endon (1; *Endone*), quietiapine (1; *quetiapine*), rydodeg (1; *Ryzodeg*), and sodabic (1; *Sodibic*).

Regarding the processing time for both models, Model 1 processed discharge summaries at a mean of 4.9 seconds per summary (SD: 4.1), while Model 2 processed candidate sentences at a mean of 3.5 seconds per sentence (SD: 0.6). Given an average of 2.2 candidate sentences per discharge summary, the combined processing time was 12.6 seconds per summary.

## 4 Discussion

This study developed and validated a two-stage hybrid approach combining rule-based NLP (Model 1) and LLM (Model 2) for automated extraction of deprescribing recommendations from discharge summaries, achieving an F1 score of 0.91 and accuracy of 0.90 in the test set. The few-shot learning strategy enabled high classification performance without parameter tuning, demonstrating the feasibility of LLM-based automated information extraction for medication management at hospital discharge.

### 4.1 Effectiveness of the two-stage hybrid approach

While rule-based approaches offer high reproducibility, they have limitations in handling diverse clinical descriptions and context-dependent reasoning.[24, 25] Recent studies have adopted similar two-stage NLP+LLM approaches using GPT-4-Turbo, including spinal surgery data extraction[26] and identification of deprescribing opportunities in emergency departments.[15] These studies share our hybrid method of leveraging rule-based precision and LLM contextual understanding.

In contrast to previous studies using GPT-4-Turbo, our use of Llama 3.3 (70B) in a local environment offers advantages in cost efficiency, data privacy compliance, and model transparency.[27-29] Particularly for large-scale implementation, the use of locally executable LLMs enables sustainable operation in terms of both cost and governance.

### 4.2 Performance characteristics and variability

The 3 most frequently used pre-defined keywords for deprescribing recommendation (i.e., “stop,” “cease,” and “wean”) accounted for 83.3% of all deprescribing recommendations, demonstrating a relatively concentrated terminology pattern in discharge summaries. This high degree of consistency validates the feasibility of our rule-based keyword approach in Model 1. The typo correction algorithm and fuzzy matching function successfully accommodated real-world documentation variability, including spelling errors and abbreviations. The distribution of deprescribing recommendation categories differed by medication type. Short-term medications such as antibiotics were predominantly recommended for immediate cessation, reflecting completion of treatment courses. In contrast, medications requiring gradual withdrawal, such as opioids, were more frequently recommended for dose reduction with aim to cease, consistent with clinical guidelines for safe opioid tapering.[30]

The inter-rater reliability for establishing the gold standard (Cohen’s kappa = 0.704) demonstrated substantial agreement.[31] However, agreement rates were different across categories, ranging from 52.4% for “dose reduction without aim to cease” to 97.8% for “cessation.” This variability reflects the subjectivity in interpreting clinical recommendations, particularly for gradual tapering strategies that may lack explicit timelines or endpoints. The lower agreement for ambiguous categories underscores a limitation in establishing ground truth and suggests the need for more explicit documentation standards in clinical practice. Substantial variation in F1 scores across hospitals (range: 0.67–0.97) may reflect differences in sample sizes and documentation styles, underscoring the importance of multi-site validation with larger samples. The consistency-based confidence analysis demonstrated excellent prediction stability, with 98.5% of cases showing perfect agreement across five independent samplings. However, calibration analysis revealed moderate miscalibration (Expected Calibration Error = 0.090), with accuracy varying non-linearly across self-reported confidence levels. The non-linear accuracy pattern demonstrates that the confidence scores do not reliably reflect prediction quality, suggesting that a threshold-based approach, where predictions above a certain confidence cutoff are automated while those below require human review, would not be feasible with the current model configuration.

Error analysis revealed that false positive errors most frequently resulted from misidentifying past actions as recommendations, specifically failing to distinguish between completed actions during hospitalisation and future recommendations on discharge. The need for such differentiation may vary between applications of the model. False negative errors were entirely due to context/meaning misunderstanding. These errors may be attributed to the line-by-line analysis approach (processing each paragraph element separated by line breaks) employed in this study, which limits access to temporal and contextual information across adjacent lines. However, the focused input approach may have contributed to the high consistency rate (98.5% perfect agreement across five samplings), suggesting a trade-off between contextual understanding and prediction stability.

### 4.3 Limitations

This study has several limitations. First, data from six hospitals in NSW, Australia may limit generalisability to other healthcare systems. Second, the rule-based approach relies on predefined keywords, potentially missing novel expressions or institution-specific terminology. Third, this study defined deprescribing to include dose reduction without aim to cease and cessation of short-term therapy (e.g., antibiotics), though these definitions remain debated in the literature.[22]

## 5 Conclusion

This study demonstrated that a two-stage hybrid approach combining rule-based NLP and open-source LLM can accurately extract deprescribing recommendations from discharge summaries, achieving an F1 score of 0.907. The use of an open-source LLM enables cost-efficient, privacy-compliant local deployment in healthcare institutions. This approach enables systematic identification of deprescribing recommendations communicated in discharge summaries, potentially facilitating hospital-GP collaboration in medication management during care transitions and evaluation of quality improvement initiatives or research.

## Acknowledgements

The authors acknowledge assistance with data collection from Stephen Mattes, Roshan Buhary, and Aiyyan Mansuri. We also acknowledge Nashwa Masnoon’s contribution to creating a list of pre-defined keywords for deprescribing, and thank Ann Mirapuri for her help and support with this project.

## Declarations

### Ethics approval

Ethics approval was granted by the Northern Sydney Local Health District Human Research Ethics Committee (2021/ETH11776).

### Informed consent

A waiver of individual patient consent was granted on the basis of the large sample size and retrospective extraction of routinely collected electronic medical record data.

### Data availability

The data underlying this article will be shared by the corresponding author upon reasonable request.

### Authors’ contributions

**Kenji Fujita:** Conceptualization; methodology; data curation; formal analysis; software; validation; visualization; writing – original draft preparation; writing – review & editing. **Marion Matheson**: Methodology; formal analysis; validation; writing – review & editing. **Bhavya Valecha**: Methodology; formal analysis; writing – review & editing. **Sarah N Hilmer**: Project administration; conceptualization; supervision; methodology; writing – review & editing; funding acquisition; resources.

