## Supplementary File for "Development and Validation of a Two-Stage NLP-LLM System for Automated Extraction of Deprescribing Recommendations from Discharge Summaries"

### TRIPOD-LLM Checklist

**Research design categories:** LLM evaluation (E), LLM methods (M)

**LLM task categories:** Classification (C)

*Reference: Gallifant J, et al. The TRIPOD-LLM reporting guideline for studies using large language models. Nat Med. 2025;31(1):60–69.*

| Section | Item | Checklist Item | Research Design | LLM Task | Page / Location in Manuscript |
|---|---|---|---|---|---|
| **Title** | **1** | Identify the study as developing, fine-tuning, and/or evaluating the performance of an LLM, specifying the task, the target population, and the outcome to be predicted. | All | All | Title page |
| **Abstract** | **2** | See TRIPOD-LLM for Abstracts. | All | All | Abstract (page 2) |
| **Background** | **3a** | Explain the healthcare context / use case (e.g., administrative, diagnostic, therapeutic, clinical workflow) and rationale for developing or evaluating the LLM, including references to existing approaches and models. | All | All | Introduction (paragraphs 1–3) |
| | **3b** | Describe the target population and the intended use of the LLM in the context of the care pathway, including its intended users in current gold standard practices (e.g., healthcare professionals, patients, public, or administrators). | E H | All | Introduction (paragraph 2) |
| **Objectives** | **4** | Specify the study objectives, including whether the study describes the initial development, fine-tuning, or validation of an LLM (or multiple stages). | All | All | Introduction (final paragraph) |
| **Data** | **5a** | Describe the sources of data separately for the training, tuning, and/or evaluation datasets and the rationale for using these data (e.g., web corpora, clinical research/trial data, EHR data, or unknown). | All | All | Methods (study design) |
| | **5b** | Describe the relevant data points and provide a quantitative and qualitative description of their distribution and other relevant descriptors of the dataset (e.g., source, languages, countries of origin). | All | All | Table 1 |
| | **5c** | Specifically state the date of the oldest and newest item of text used in the development process (training, fine-tuning, reward modeling) and in the evaluation datasets. | All | All | Methods (study design) |
| | **5d** | Describe any data pre-processing and quality checking, including whether this was similar across text corpora, institutions, and relevant socio-demographic groups. | All | All | Methods (model 1) |
| | **5e** | Describe how missing and imbalanced data were handled and provide reasons for omitting any data. | All | All | Methods (study design) |
| **Analytical Methods** | **6a** | Report the LLM name, version, and last date of training. | All | All | Methods (model 2) |
| | **6b** | Report details of LLM development process, such as LLM architecture, training, fine-tuning procedures, and alignment strategy (e.g., reinforcement learning, direct preference optimization, etc.) and alignment goals. | M D | All | N/A<br>Study used an existing pre-trained model without fine-tuning. |
| | **6c** | Report details of how text was generated using the LLM, including any prompt engineering (including consistency of outputs), and inference settings (e.g., seed, temperature, max token length, penalties), as relevant. | M D E | All | Methods (model 2)<br>Full prompt in Supplementary File 2. |
| | **6d** | Specify the initial and post-processed output of the LLM (e.g., probabilities, classification, unstructured text). | All | All | Methods (model 2) |

| | | | | | |
|---|---|---|---|---|---|
| | **6e** | Provide details and rationale for any classification and, if applicable, how the probabilities were determined and thresholds identified. | All | C OF | Methods (model 2) |
| **LLM Output** | **7a** | Include metrics that capture the quality of generative outputs, such as consistency, relevance, and accuracy, compared to gold standards. | All | QA IR DG SS MT | N/A |
| | **7b** | Report the outcome metrics' relevance to downstream task at deployment time and, where applicable, correlation of metric to human evaluation of the text for the intended use. | E H | All | Results and Discussion |
| | **7c** | Clearly define the outcome, how the LLM predictions were calculated (e.g., formula, code, object, API), the date of inference for closed-source LLMs, and evaluation metrics. | E H | All | Methods (model 2 and evaluation) |
| | **7d** | If outcome assessment requires subjective interpretation, describe the qualifications of the assessors, any instructions provided, relevant information on demographics of the assessors, and inter-assessor agreement. | All | All | Methods (gold standard) |
| | **7e** | Specify how performance was compared to other LLMs, humans, and other benchmarks or standards. | All | All | Discussion |
| **Annotation** | **8a** | If annotation was done, report how text was labeled, including providing specific annotation guidelines with examples. | All | All | Methods (gold standard) |
| | **8b** | If annotation was done, report how many annotators labeled the dataset(s), including the proportion of data in each dataset that were annotated by more than 1 annotator, and the inter-annotator agreement. | All | All | Methods (gold standard) |
| | **8c** | If annotation was done, provide information on the background and experience of the annotators or characteristics of any models involved in labelling. | All | All | Methods (gold standard) |
| **Prompting** | **9a** | If research involved prompting LLMs, provide details on the processes used during prompt design, curation, and selection. | All | All | Methods (model 2)<br>Full prompt in Supplementary File 2. |
| | **9b** | If research involved prompting LLMs, report what data were used to develop the prompts. | All | All | Methods (model 2) |
| **Summarization** | **10** | Describe any preprocessing of the data before summarization. | All | SS | N/A |
| **Instruction tuning / Alignment** | **11** | If instruction tuning/alignment strategies were used, what were the instructions, data, and interface used for evaluation, and what were the characteristics of the populations doing evaluation? | M D | All | N/A |
| **Compute** | **12** | Report compute, or proxies thereof (e.g., time on what and how many machines, cost on what and how many machines, inference time, FLOPs), required to carry out methods. | M D E | All | Results ([processing time] |
| **Ethical Approval** | **13** | Name the institutional research board or ethics committee that approved the study and describe the participant-informed consent or the ethics committee waiver of informed consent. | All | All | Methods (study design) |
| **Open Science** | **14a** | Give the source of funding and the role of the funders for the present study. | All | All | Funding |
| | **14b** | Declare any conflicts of interest and financial disclosures for all authors. | All | All | Declaration of competing interest |
| | **14c** | Indicate where the study protocol can be accessed or state that a protocol was not prepared. | H | All | N/A |
| | **14d** | Provide registration information for the study, including register name and registration number, or state that the study was not registered. | H | All | N/A |
| | **14e** | Provide details of the availability of the study data. | All | All | Data availability |
| | **14f** | Provide details of the availability of the code to reproduce the study results. | All | All | Prompt template in Supplementary File 2. |

| | | | | | |
|---|---|---|---|---|---|
| **Public Involvement** | **15** | Provide details of any patient and public involvement during the design, conduct, reporting, interpretation, or dissemination of the study or state no involvement. | H | All | N/A |
| **Participants** | **16a** | When using patient/EHR data, describe the flow of text/EHR/patient data through the study, including the number of documents/questions/participants with and without the outcome/label and follow-up time as applicable. | E H | All | Figure 1<br>Tables 2–3. |
| | **16b** | When using patient/EHR data, report the characteristics overall and, for each data source or setting, and for development/evaluation splits, including the key dates, key characteristics, and sample size. | E H | All | Table 1 |
| | **16c** | For LLM evaluation that include clinical outcomes, show a comparison of the distribution of important clinical variables that may be associated with the outcome between development and evaluation data, if available. | E H | All | Table 1 |
| | **16d** | When using patient/EHR data, specify the number of participants and outcome events in each analysis (e.g., for LLM development, hyperparameter tuning, LLM evaluation). | E H | All | Results<br>Tables 2–3. |
| **Performance** | **17** | Report LLM performance according to pre-specified metrics (see item 7a) and/or human evaluation (see item 7d). | All | All | Table 2<br>Table 3<br>Figure 2 |
| **LLM Updating** | **18** | If applicable, report the results from any LLM updating, including the updated LLM and subsequent performance. | All | All | N/A |
| **Interpretation** | **19a** | Give an overall interpretation of the main results, including issues of fairness in the context of the objectives and previous studies. | All | All | Discussion (principal findings) |
| **Limitations** | **19b** | Discuss any limitations of the study and their effects on any biases, statistical uncertainty, and generalizability. | All | All | Limitations |
| **Usability of the LLM in context** | **19c** | Describe any known challenges in using data for the specified task and domain context with reference to representation, missingness, harmonization, and bias. | E H | All | Discussion (e.g., documentation variability across hospitals) |
| | **19d** | Define the intended use for the implementation under evaluation, including the intended input, end-user, level of autonomy/human oversight. | E H | All | Discussion/Conclusion |
| | **19e** | If applicable, describe how poor quality or unavailable input data should be assessed and handled when implementing the LLM. | E H | All | Discussion: Typo correction and fuzzy matching address poor quality input. Line-by-line processing trade-off discussed. |
| | **19f** | If applicable, specify whether users will be required to interact in the handling of the input data or use of the LLM, and what level of expertise is required of users. | E H | All | Discussion: Human review required for low-confidence predictions; clinical expertise needed for ambiguous cases. |
| | **19g** | Discuss any next steps for future research, with a specific view to applicability and generalizability of the LLM. | All | All | Conclusion |

LLM = large language model; M = LLM methods; D = de novo LLM development; E = LLM evaluation; H = LLM evaluation in healthcare settings; C = classification; OF = outcome forecasting; QA = long-form question-answering; IR = information retrieval; DG = document generation; SS = summarization and simplification; MT = machine translation; EHR = electronic health record.

Note: For studies using existing LLMs, users should include reference(s) to reportable information if provided by the original developers or state that this information is not available.

Supplementary File 1. List of pre-defined keywords for deprescribing recommendation

weaning, wean, be weaned,
ceasing, cease, be ceased, cessation,
deprescribing, deprescribe, be deprescribed, deprescription
de-prescribing, de-prescribe, be de-prescribed, de prescription
titrating, titrate, be titrated, titration,
down-titrating, down-titrate, be down-titrated, down-titration,
withholding, withhold, be withheld,
rationalising, rationalise, be rationalised,
stepping down, step down, be stepped down,
stopping, stop, be stopped,
reducing, reduce, be reduced, reduction
decreasing, decrease, be decreased,
rotating, rotate, be rotated,
half, be halved,
modifying, modify, be modified, modification,
adjusting, adjust, be adjusted, adjustment,
changing, change, be changed,
switching, switch, be switched,
ongoing use, ongoing need, ongoing requirement,
alteration, alternative,

Supplementary File 2. Final version of the deprescribing classification prompt

Analyze the following text for medication recommendations related to dose reduction or cessation:
"[SENTENCE]"

Consider this targeted medication: [MEDICATION]

For the targeted medication, classify the recommendation into one of these categories:

1. No: No recommendation for dose reduction or cessation. (Not a deprescribing recommendation)
   - Includes:
     - Maintaining the same dose
     - Increasing (up-titrating) the dose
     - General advice unrelated to dose (e.g., re-check labs, monitor)
     - Starting a new medication without changing the targeted medication
     - Switching from one medication to another (e.g., "please change drug A to drug B")
     - Changing the route of administration (e.g., "step down from IV drug A to oral drug A")
     - Medication was already ceased or had its dose reduced before discharge (e.g., "*** CHANGE *** Rosuvastatin reduced to 20 mg daily")
   - Examples:
     - "Smoking cessation encouraged"

2. Dose adjust (Unclear if up or down titration): Recommendation to adjust or titrate the dose, but not clearly stated as increase or decrease.
   - Includes:
     - Using words like "titrate" or "adjust" without specifying direction
     - Instructions to modify dose based on patient condition or test results, with no clear up/down
   - Examples:
     - "Kindly recheck INR and titrate Warfarin accordingly."
     - "Review fluid status and titrate Frusemide as needed."

3. Dose reduction without aim to cease: Clear recommendation to reduce the dose, but not to stop the medication entirely, or it is unclear whether there is an eventual intention to discontinue.
   - Includes:
     - Using words like "reduce," "decrease," or "step down"
     - Switching to a lower maintenance dose
     - Lowering frequency from multiple times/day to fewer
   - Examples:
     - "ADDED Pantoprazole 40 mg twice daily for 1 month, then step down to daily dosing."
     - "Started on cholecalciferol 125 microg daily for 1 month, then reduce to 25–50 microg daily for maintenance."
     - "PO amiodarone 200 mg BD for total 1 week, then decrease to 200 mg daily."

4. Dose reduction with aim to cease: Recommendation to reduce the dose with the goal or plan of ultimately stopping it.
   - Includes:
     - Using words like "wean" or "taper" with intent to discontinue

    - Phrases such as "with aim to cease" or "with the goal to cease"
    - Instructions to move to PRN (as needed) dosing
    - Any mention of gradual dose reduction specifically leading to cessation
  - Examples:
    - "GP to please review Endone and Targin use and wean as appropriate."
    - "Aim to reduce this to PRN dosing after 1 week of discharge."

5. Cease: Direct or definite recommendation to stop the medication.
  - Includes:
    - Using words like "cease," "stop," or "discontinue"
    - Specifying a stop date or plan to stop completely
    - Time-limited prescriptions (e.g., "take for 7 days then stop")
  - Examples:
    - (targeted medication is Targin) "Please start Paracetamol 1000 mg TDS following cessation of Targin therapy."
    - (targeted medication is sitagliptin) "Once recovered from pyelonephritis, GP can consider ozempic therapy with downtitration of insulin and cessation of sitagliptin."

Important notes:
  - Ignore recommendations for any medications other than the targeted medication.
  - If the targeted medication is not explicitly mentioned or there's no clear recommendation for it, classify as category 1 (No).
  - If the text is simply reporting that the medication was already reduced or ceased during admission (i.e., not a deprescribing recommendation), it should be classified as category 1.
  - Do not include reasoning in the output—just the category.

Answer format:
  "[MEDICATION NAME], Category: [CATEGORY NUMBER]"

Provide only one answer for the targeted medication.

Answer:

Supplementary File 3. Model 2 performance by hospital (training and test set)

Training dataset

| Hospital | Sensitivity | Specificity | PPV | NPV | F1 Score | Accuracy |
|---|---|---|---|---|---|---|
| Hospital A (n=122) | 0.91 | 0.87 | 0.90 | 0.89 | 0.90 | 0.89 |
| Hospital B (n=110) | 0.98 | 0.77 | 0.85 | 0.97 | 0.91 | 0.89 |
| Hospital C (n=118) | 0.97 | 0.87 | 0.90 | 0.96 | 0.93 | 0.92 |
| Hospital D (n=129) | 1.00 | 0.83 | 0.83 | 1.00 | 0.91 | 0.91 |
| Hospital E (n=26) | 1.00 | 0.54 | 0.68 | 1.00 | 0.81 | 0.77 |
| Hospital F (n=132) | 1.00 | 0.79 | 0.86 | 1.00 | 0.93 | 0.91 |
| Total (n=637) | 0.97 | 0.81 | 0.86 | 0.96 | 0.91 | 0.90 |

Test dataset

| Hospital | Sensitivity | Specificity | PPV | NPV | F1 Score | Accuracy |
|---|---|---|---|---|---|---|
| Hospital A (n=12) | 0.63 | 0.50 | 0.71 | 0.40 | 0.67 | 0.58 |
| Hospital B (n=35) | 0.95 | 0.93 | 0.95 | 0.93 | 0.95 | 0.94 |
| Hospital C (n=34) | 1.00 | 0.81 | 0.86 | 1.00 | 0.92 | 0.91 |
| Hospital D (n=27) | 1.00 | 0.92 | 0.93 | 1.00 | 0.97 | 0.96 |
| Hospital E (n=9) | 1.00 | 0.50 | 0.50 | 1.00 | 0.67 | 0.67 |
| Hospital F (n=27) | 1.00 | 0.92 | 0.93 | 1.00 | 0.97 | 0.96 |
| Total (n=144) | 0.95 | 0.84 | 0.87 | 0.93 | 0.91 | 0.90 |

Abbreviation, PPV: positive predictive value, NPV: negative predictive valu

Supplementary File 4. Confusion matrices for five-category deprescribing recommendation classification at sentence level (n=1,061)

| | | Gold Standard | | | | |
|---|---|---|---|---|---|---|
| | | Cessation | Dose reduction with aim to cease | Dose reduction without aim to cease | Dose adjust (unclear) | No deprescribing |
| LLM | Cessation | 325 | 35 | 5 | 0 | 3 |
| | Dose reduction with aim to cease | 5 | 162 | 17 | 7 | 3 |
| | Dose reduction without aim to cease | 0 | 11 | 27 | 1 | 0 |
| | Dose adjust (unclear) | 0 | 2 | 1 | 164 | 11 |
| | No deprescribing | 34 | 25 | 25 | 9 | 189 |

Supplementary File 5. The 20 most frequently recommended medications for deprescribing

| No. | Active Ingredient Finalised | Cease | Dose reduction with aim to cease | Dose reduction without aim to cease | Total |
|---|---|---|---|---|---|
| 1 | amoxicillin (incl. amoxicillin-clavulanate) | 50 (100%) | 0 | 0 | 50 (12%) |
| 2 | oxycodone +/- naloxone | 13 (31%) | 29 (69%) | 0 | 42 (10%) |
| 3 | prednisolone | 22 (63%) | 12 (34%) | 1 (3%) | 35 (8.4%) |
| 4 | cefalexin | 19 (100%) | 0 | 0 | 19 (4.5%) |
| 5 | paracetamol | 3 (18%) | 14 (82%) | 0 | 17 (4.1%) |
| 6 | dexamethasone | 10 (71%) | 4 (29%) | 0 | 14 (3.3%) |
| 7 | ciprofloxacin | 10 (100%) | 0 | 0 | 10 (2.4%) |
| 8 | clopidogrel | 10 (100%) | 0 | 0 | 10 (2.4%) |
| 9 | aspirin | 8 (100%) | 0 | 0 | 8 (1.9%) |
| 10 | morphine | 1 (13%) | 7 (88%) | 0 | 8 (1.9%) |
| 11 | pantoprazole | 5 (63%) | 1 (13%) | 2 (25%) | 8 (1.9%) |
| 12 | tapentadol | 3 (38%) | 5 (63%) | 0 | 8 (1.9%) |
| 13 | doxycycline | 7 (100%) | 0 | 0 | 7 (1.7%) |
| 14 | metronidazole | 7 (100%) | 0 | 0 | 7 (1.7%) |
| 15 | cefuroxime | 6 (100%) | 0 | 0 | 6 (1.4%) |
| 16 | colecalciferol | 1 (17%) | 1 (17%) | 4 (67%) | 6 (1.4%) |
| 17 | macrogol 3350 | 4 (67%) | 2 (33%) | 0 | 6 (1.4%) |
| 18 | pregabalin | 0 | 4 (67%) | 2 (33%) | 6 (1.4%) |
| 19 | amiodarone | 3 (60%) | 0 | 2 (40%) | 5 (1.2%) |
| 20 | apixaban | 3 (60%) | 2 (40%) | 0 | 5 (1.2%) |
